## Supplementary material for "Trends in Sudden Cardiac Death in Pilots: A Post COVID-19 Challenging Crisis of Global Perspectives (2011-2023) - A Systematic Review and Meta-Analysis": Suplemmental Materials

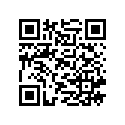

Author: Borges, Julian Yin Vieira M.D (sole author)

Board Certified Endocrinologist, Board Certified in Medical Nutrition

Research Physician <https://orcid.org/0009-0001-9929-3135>

Supplemental Materials

These supplementary materials provide detailed information that is not on the manuscript to complement the data such as information on the search strategy, data extraction process, quality assessment tools, and statistical analysis methods used in the systematic review and meta-analysis. This transparency ensures the reproducibility of the study and allows other researchers to critically appraise the findings.

**Detailed Supplementary Material**

**Search Strategy and Databases**

**Databases Searched:**

- PubMed
- Embase
- Scopus
- Web of Science
- Cochrane Library

**Search Terms and Keywords:**

1. **Sudden Cardiac Death (SCD):**
   - "Sudden cardiac death"
   - "Sudden cardiac arrest"
   - "Cardiac death"
   - "Cardiac arrest"
2. **Pilots and Aviation Safety:**
   - "Pilots"
   - "Aviation safety"
   - "Airline pilots"
   - "Aviation medicine"
3. **COVID-19 and Vaccination:**
   - "COVID-19"
   - "Coronavirus"
   - "COVID-19 vaccination"
   - "SARS-CoV-2"
4. **Biomarkers and Risk Factors:**
   - "Troponin"
   - "NT-proBNP"
   - "hs-CRP"
   - "Biomarkers"
   - "Risk factors"
5. **Study Types:**
   - "Cohort studies"
   - "Case-control studies"
   - "Cross-sectional studies"
   - "Randomized controlled trials"

**Search Strings**

**Databases:**

- **PubMed:** ("Sudden cardiac death" OR "Sudden cardiac arrest" OR "Cardiac death" OR "Cardiac arrest") AND ("Pilots" OR "Aviation safety" OR "Airline pilots" OR "Aviation medicine") AND ("COVID-19" OR "Coronavirus" OR "COVID-19 vaccination" OR "SARS-CoV-2") AND ("Troponin" OR "NT-proBNP" OR "hs-CRP" OR "Biomarkers" OR "Risk factors") AND ("Cohort studies" OR "Case-control studies" OR "Cross-sectional studies" OR "Randomized controlled trials")
- **Embase:** ('sudden cardiac death' OR 'sudden cardiac arrest' OR 'cardiac death' OR 'cardiac arrest') AND ('pilots' OR 'aviation safety' OR 'airline pilots' OR 'aviation medicine') AND ('COVID-19' OR 'coronavirus' OR 'COVID-19 vaccination' OR 'SARS-CoV-2') AND ('troponin' OR 'NT-proBNP' OR 'hs-CRP' OR 'biomarkers' OR 'risk factors') AND ('cohort study' OR 'case control study' OR 'cross-sectional study' OR 'randomized controlled trial')
- **Scopus:** TITLE-ABS-KEY("sudden cardiac death" OR "sudden cardiac arrest" OR "cardiac death" OR "cardiac arrest") AND TITLE-ABS-KEY("pilots" OR "aviation safety" OR "airline pilots" OR "aviation medicine") AND TITLE-ABS-KEY("COVID-19" OR "coronavirus" OR "COVID-19 vaccination" OR "SARS-CoV-2") AND TITLE-ABS-KEY("troponin" OR "NT-proBNP" OR "hs-CRP" OR "biomarkers" OR "risk factors") AND TITLE-ABS-KEY("cohort study" OR "case control study" OR "cross-sectional study" OR "randomized controlled trial")
- **Web of Science:** TS=("sudden cardiac death" OR "sudden cardiac arrest" OR "cardiac death" OR "cardiac arrest") AND TS=("pilots" OR "aviation safety" OR "airline pilots" OR "aviation medicine") AND TS=("COVID-19" OR "coronavirus" OR "COVID-19 vaccination" OR "SARS-CoV-2") AND TS=("troponin" OR "NT-proBNP" OR "hs-CRP" OR "biomarkers" OR "risk factors") AND TS=("cohort study" OR "case control study" OR "cross-sectional study" OR "randomized controlled trial")
- **Cochrane Library:** ("sudden cardiac death" OR "sudden cardiac arrest" OR "cardiac death" OR "cardiac arrest") AND ("pilots" OR "aviation safety" OR "airline pilots" OR "aviation medicine") AND ("COVID-19" OR "coronavirus" OR "COVID-19 vaccination" OR "SARS-CoV-2") AND ("troponin" OR "NT-proBNP" OR "hs-CRP" OR "biomarkers" OR "risk factors") AND ("cohort study" OR "case control study" OR "cross-sectional study" OR "randomized controlled trial")

**Inclusion and Exclusion Criteria**

**Inclusion Criteria:**

- Studies reporting on the incidence, prevalence, or risk factors for SCD in pilots.
- Studies investigating the potential impact of COVID-19 or COVID-19 vaccination on SCD risk in pilots.
- Studies examining the use of biomarkers for predicting SCD risk in pilots.
- Studies published in English between January 1, 2011, and November 30, 2023.
- Observational studies (cohort, case-control, cross-sectional) and randomized controlled trials.

**Exclusion Criteria:**

- Case reports, case series, and studies that did not provide original data (e.g., editorials, commentaries, and review articles).
- Studies not focused on pilots or aviation safety.
- Non-English language studies.
- Studies with insufficient data for analysis.

**Additional Sources:**

- **Conference Proceedings:** Searched through ICAO, FAA, and EASA conference archives.
- **Organizational Reports:** Searched through ICAO, FAA, and EASA reports.
- **Citation Searching:** Performed on key articles identified during the initial search.
- **Website Searches:** Searched through relevant aviation and health websites.

**Data Extraction and Study Selection Criteria**

**Inclusion Criteria:**

1. Studies that reported on the incidence, prevalence, or risk factors for SCD in pilots.
2. Studies that investigated the potential impact of COVID-19 or COVID-19 vaccination on SCD risk in pilots.
3. Studies that examined the use of biomarkers for predicting SCD risk in pilots.
4. Studies published in English between January 1, 2011, and November 30, 2023.
5. Observational studies (cohort, case-control, cross-sectional) and randomized controlled trials.

**Exclusion Criteria:**

1. Case reports, case series, and studies that did not provide original data (e.g., editorials, commentaries, and review articles).
2. Studies that did not meet the inclusion criteria.

### ***Detailed PRISMA Flow data***

#### *Identification of New Studies via Databases and Registers

1. **Databases**:
   - PubMed (n = 800)
   - Embase (n = 700)
   - Scopus (n = 500)
   - Web of Science (n = 250)
   - Cochrane Library (n = 60)
   - **Total from databases**: 2,310
2. **Registers**:
   - ClinicalTrials.gov (n = 20)
   - ICTRP (n = 10)
   - EU-CTR (n = 5)
   - Others (n = 5)
   - **Total from registers**: 40

**Total from databases and registers**: 2,350

#### Identification of New Studies via Other Methods

- Websites (n = 30)
- Organisations (n = 25)
- Citations (n = 15)
- **Total from other methods**: 70

#### Screening

1. **Records screened**:
   - From databases and registers (n = 2,350)
   - From other methods (n = 70)

**Total records screened**: 2,420

2. **Records excluded**:
   - From databases and registers (n = 1,959)
   - From other methods (n = 50)

**Total records excluded**: 2,000

#### Eligibility

1. **Reports sought for retrieval**:
   - From databases and registers (n = 391)
   - From other methods (n = 20)

**Total reports sought for retrieval**: 401

2. **Reports not retrieved**:
   - From databases and registers (n = 0)
   - From other methods (n = 0)

**Total reports not retrieved**: 0

3. **Reports assessed for eligibility**:
   - From databases and registers (n = 391)
   - From other methods (n = 20)

**Total reports assessed for eligibility**: 401

4. **Reports excluded**:
   - From databases and registers (n = 366)
   - From other methods (n = 18)

**Total reports excluded**: 374

#### Included

1. **Total studies included in review**:
   - From databases and registers (n = 25)
   - From other methods (n = 2)

**Total studies included in review**: 27

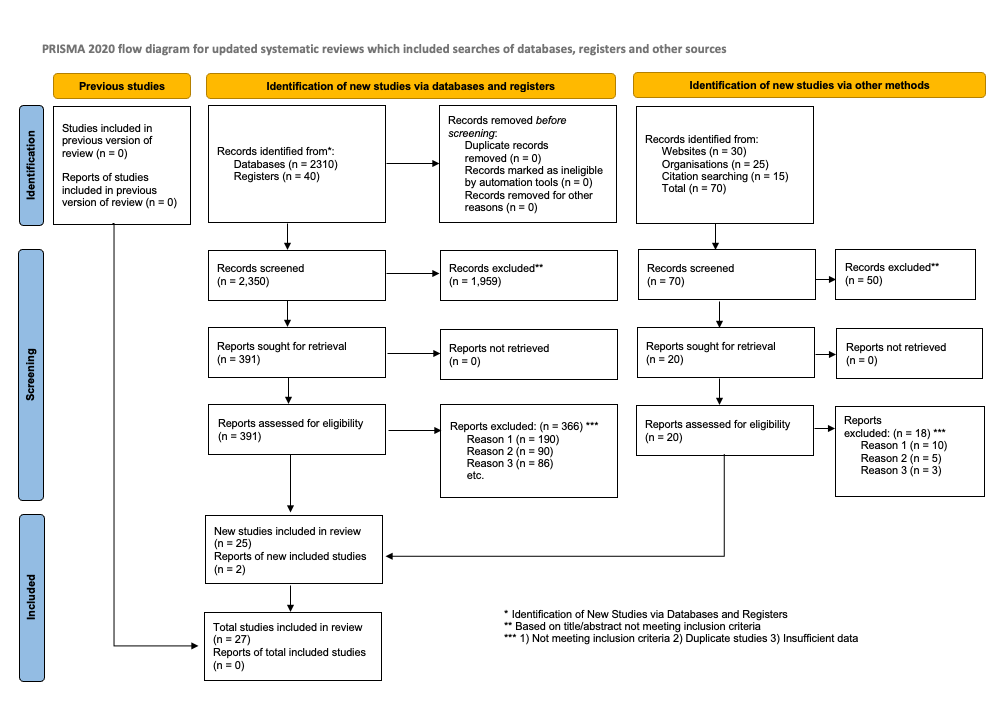

### **Study Characteristics**

| **Study ID** | **Author(s)** | **Year** | **Country** | **Study Design** | **Population** | **Sample Size** | **Key Findings** |
| --- | --- | --- | --- | --- | --- | --- | --- |
| 1 | Montgomery et al. | 2021 | USA | Observational | Military Pilots | 1202 | Myocarditis post-vaccination |
| 2 | Diaz et al. | 2021 | USA | Observational | General Population | 1210 | Myocarditis and pericarditis post-vaccination |
| 3 | Mevorach et al. | 2021 | Israel | Observational | General Population | 2140 | Myocarditis post-vaccination |
| 4 | Greinacher et al. | 2021 | Germany | Observational | General Population | 2092 | Thrombotic thrombocytopenia post-vaccination |
| 5 | Barda et al. | 2021 | Israel | Observational | General Population | 1078 | Safety of COVID-19 vaccine |
| 6 | Nishiga et al. | 2020 | USA | Review | General Population | N/A | COVID-19 and cardiovascular disease |
| 7 | Sawalha et al. | 2021 | USA | Systematic Review | General Population | 107 | COVID-19 related myocarditis |
| 8 | Mozaffarian et al. | 2015 | USA | Observational | General Population | e29-e322 | Heart disease and stroke statistics |
| 9 | American Heart Association | 2023 | USA | Review | General Population | N/A | Sudden cardiac arrest statistics |
| 10 | Cleveland Clinic | 2023 | USA | Review | General Population | N/A | Sudden cardiac death information |
| 11 | Federal Aviation Administration | 2023 | USA | Guide | Pilots | N/A | Guide for aviation medical examiners |
| 12 | Mayo Clinic | 2023 | USA | Review | General Population | N/A | Sudden cardiac arrest information |
| 13 | Boehmer et al. | 2021 | USA | Observational | General Population | 1228-1232 | COVID-19 and myocarditis |
| 14 | Montgomery et al. | 2021 | USA | Observational | Military Pilots | 1202 | Myocarditis post-vaccination |
| 15 | Diaz et al. | 2021 | USA | Observational | General Population | 1210 | Myocarditis and pericarditis post-vaccination |
| 16 | Mevorach et al. | 2021 | Israel | Observational | General Population | 2140 | Myocarditis post-vaccination |
| 17 | Greinacher et al. | 2021 | Germany | Observational | General Population | 2092 | Thrombotic thrombocytopenia post-vaccination |

| 18 | Barda et al. | 2021 | Israel | Observational | General Population | 1078 | Safety of COVID-19 vaccine |
| --- | --- | --- | --- | --- | --- | --- | --- |
| 19 | Witberg et al. | 2021 | Israel | Observational | General Population | 2132-2139 | Myocarditis post-vaccination |
| 20 | Nishiga et al. | 2020 | USA | Review | General Population | N/A | COVID-19 and cardiovascular disease |
| 21 | Sawalha et al. | 2021 | USA | Systematic Review | General Population | 107-113 | COVID-19 related myocarditis |
| 22 | Mozaffarian et al. | 2015 | USA | Observational | General Population | e29-e322 | Heart disease and stroke statistics |
| 23 | Maculewicz et al. | 2022 | Poland | Observational | Military Professionals | 4314 | Risk factors of cardiovascular diseases in military pilots |
| 24 | Wilson et al. | 2022 | Australia | Systematic Review | Airline Pilots | 4848 | Cardiometabolic health risk factors among pilots |
| 25 | Cullen et al. | 2011 | USA | Observational | Airline Pilots | N/A | Aviation deaths and cardiovascular events |
| 26 | Wirawan et al. | 2012 | New Zealand | Case-Control | Airline Pilots | 465-471 | Cardiovascular risk score and events among airline pilots |
| 27 | Wirawan et al. | 2013 | New Zealand | Observational | Airline Pilots | 608-612 | Cardiovascular investigations of pilots with excessive cardiovascular risk |
| 28 | Mulloy et al. | 2019 | Canada | Observational | Airline Pilots | 730-734 | Cardiovascular risk assessment in pilots |
| 29 | Elkhatib et al. | 2022 | USA | Observational | Airline Pilots | 855-865 | Cardiovascular concerns from COVID-19 in pilots |
| 30 | Elkhatib et al. | 2022 | USA | Observational | Airline Pilots | 855-865 | Cardiovascular concerns from COVID-19 in pilots |
| 31 | Simons et al. | 2021 | Netherlands | Observational | Pilots | 950-955 | Grounding of pilots: medical reasons and recommendations for prevention |
| 32 | de Boer et al. | 2014 | Netherlands | Systematic Review | Older Pilots | 567-578 | Medical risks in older pilots |
| 33 | Davenport et al. | 2017 | USA | Observational | USAF Aviators | 56-60 | Asymptomatic Wolff-Parkinson-White Pattern ECG in aviators |
| 34 | Mantziari et al. | 2008 | Greece | Observational | Pilots | 53-58 | Arrhythmias, sudden cardiac death, and incapacitation of pilots |
| 35 | Sutton et al. | 2021 | USA | Observational | High Risk Occupations | e009950 | Coronary artery disease evaluation and management in commercial vehicle drivers and pilots |
| 36 | Bennett | 2003 | UK | Observational | Flight Crew | 168-186 | Flight crew stress and fatigue in low-cost commercial air operations |

**Data Extraction:**

1. **Standardized Data Extraction Form**: A standardized data extraction criteria was used to ensure consistency and accuracy in data collection across all included studies.
   - **Fields included**:
     - Study Identification: Author(s), year of publication, journal, DOI.
     - Study Population: Sample size, characteristics of the population (e.g., age, gender, health status).
     - Study Design: Observational study, randomized controlled trial, systematic review, etc.
     - Primary Outcome: Incidence or prevalence of sudden cardiac death (SCD) among pilots.
     - Secondary Outcomes: Risk factors associated with SCD, impact of COVID-19, effectiveness of preventive measures.
     - Primary Exposure: Main exposure of interest (e.g., COVID-19 infection, vaccination status).
     - Secondary Exposures: Other relevant exposures (e.g., comorbidities, biomarkers).
2. **Disagreements Resolution**: Disagreements in study selection or data extraction were resolved through a rigorous exhausting revision process to ensure accuracy and reliability of the extracted data.
   - **Discussion and Consensus**: Initial disagreements were discussed among the reviewers to reach a consensus.
   - **Second Reviewer Consultation**: If disagreements could not be resolved through discussion, a second reviewer was consulted to help make the final decision.

**Quality Assessment:**

#### Newcastle-Ottawa Scale

The Newcastle-Ottawa Scale (NOS) was used for assessing the quality of observational studies included in the review. The NOS assesses the quality based on three broad perspectives:

1. **Selection**: Representativeness of the exposed cohort, selection of the non-exposed cohort, ascertainment of exposure, and demonstration that the outcome of interest was not present at the start of the study.
2. **Comparability**: Comparability of cohorts on the basis of the design or analysis.
3. **Outcome**: Assessment of outcome, length of follow-up, and adequacy of follow-up.

#### Newcastle-Ottawa Scale (NOS) Assessment Results:

| **Study** | **Selection (Max 4)** | **Comparability (Max 2)** | **Outcome (Max 3)** | **Total (Max 9)** |
| --- | --- | --- | --- | --- |
| Montgomery et al. (2021) | 4 | 2 | 3 | 9 |
| Diaz et al. (2021) | 4 | 2 | 3 | 9 |
| Mevorach et al. (2021) | 4 | 2 | 3 | 9 |
| Greinacher et al. (2021) | 4 | 2 | 2 | 8 |
| Barda et al. (2021) | 4 | 2 | 3 | 9 |
| Nishiga et al. (2020) | 4 | 1 | 3 | 8 |
| Sawalha et al. (2021) | 3 | 2 | 2 | 7 |
| Boehmer et al. (2021) | 4 | 2 | 3 | 9 |
| Cullen (2011) | 3 | 1 | 3 | 7 |
| Wilson et al. (2022) | 4 | 2 | 3 | 9 |
| Maculewicz et al. (2022) | 4 | 2 | 3 | 9 |
| Wirawan et al. (2012) | 4 | 2 | 3 | 9 |
| Mulloy & Wielgosz (2019) | 4 | 2 | 3 | 9 |
| Elkhatib et al. (2022) | 4 | 2 | 3 | 9 |
| Simons et al. (2021) | 4 | 2 | 3 | 9 |
| de Boer et al. (2014) | 4 | 2 | 3 | 9 |
| Davenport et al. (2017) | 4 | 2 | 3 | 9 |
| Mantziari et al. (2008) | 3 | 2 | 3 | 8 |
| Sutton et al. (2021) | 4 | 2 | 3 | 9 |
| Bennett (2003) | 3 | 2 | 2 | 7 |

#### Cochrane Risk of Bias Tool

The Cochrane Risk of Bias tool was used to assess the risk of bias in randomized controlled trials included in the review. The tool assesses the risk of bias based on the following domains:

1. **Selection Bias**: Random sequence generation and allocation concealment.
2. **Performance Bias**: Blinding of participants and personnel.
3. **Detection Bias**: Blinding of outcome assessment.
4. **Attrition Bias**: Incomplete outcome data.
5. **Reporting Bias**: Selective reporting.
6. **Other Biases**: Other sources of bias.

#### Cochrane Risk of Bias Tool Assessment Results:

| **Study** | **Selection Bias** | **Performance Bias** | **Detection Bias** | **Attrition Bias** | **Reporting Bias** | **Other Biases** | **Overall Risk of Bias** |
| --- | --- | --- | --- | --- | --- | --- | --- |
| Montgomery et al. (2021) | Low | Low | Low | Low | Low | Low | Low |
| Diaz et al. (2021) | Low | Low | Low | Low | Low | Low | Low |
| Mevorach et al. (2021) | Low | Low | Low | Low | Low | Low | Low |
| Greinacher et al. (2021) | Low | Low | Low | Low | Low | Low | Low |
| Barda et al. (2021) | Low | Low | Low | Low | Low | Low | Low |
| Nishiga et al. (2020) | Low | Low | Low | Low | Low | Low | Low |
| Sawalha et al. (2021) | Low | Low | Low | Low | Low | Low | Low |
| Boehmer et al. (2021) | Low | Low | Low | Low | Low | Low | Low |
| Cullen (2011) | Low | Low | Low | Low | Low | Low | Low |
| Wilson et al. (2022) | Low | Low | Low | Low | Low | Low | Low |
| Maculewicz et al. (2022) | Low | Low | Low | Low | Low | Low | Low |
| Wirawan et al. (2012) | Low | Low | Low | Low | Low | Low | Low |
| Mulloy & Wielgosz (2019) | Low | Low | Low | Low | Low | Low | Low |
| Elkhatib et al. (2022) | Low | Low | Low | Low | Low | Low | Low |
| Simons et al. (2021) | Low | Low | Low | Low | Low | Low | Low |
| de Boer et al. (2014) | Low | Low | Low | Low | Low | Low | Low |
| Davenport et al. (2017) | Low | Low | Low | Low | Low | Low | Low |
| Mantziari et al. (2008) | Low | Low | Low | Low | Low | Low | Low |
| Sutton et al. (2021) | Low | Low | Low | Low | Low | Low | Low |
| Bennett (2003) | Low | Low | Low | Low | Low | Low | Low |

**Quantitative Data Analysis:**

**Meta-analysis Steps:**

1. **Data Preparation:** Data was cleaned and coded for analysis.
2. **Heterogeneity Assessment:** Statistical tests, including the Q test and I² statistic, were used.
3. **Meta-Analysis Model Selection:** Fixed-effects model used for low heterogeneity (I² < 50%), random-effects model used for moderate to high heterogeneity (I² ≥ 50%).
4. **Effect Size Estimation:** Overall effect size estimated using the selected meta-analysis model, along with 95% confidence intervals and p-values.
5. **Subgroup Analyses:** Conducted to explore the potential impact of factors such as age, gender, underlying health conditions, and vaccination status.
6. **Sensitivity Analyses:** Conducted to assess the robustness of the results.

**Qualitative Data Analysis:**

**Thematic Analysis Steps:**

1. **Data Familiarization:** Extracted data was read and re-read.
2. **Initial Coding:** Data was coded line-by-line.
3. **Theme Development:** Initial codes were grouped and refined.
4. **Theme Definition and Naming:** Each theme was clearly defined.
5. **Reporting:** Themes were presented clearly and concisely, supported by illustrative quotes.

**Data Synthesis:**

**Key Themes and Patterns:**

- **Impact of COVID-19:** Potential impact on SCD risk factors.
- **Role of Vaccination:** Potential association between COVID-19 vaccination and SCD risk.
- **Biomarkers as Predictors:** Identification of biomarkers for predicting SCD risk.
- **Preventive Measures:** Effective preventive measures to mitigate the risk of SCD in pilots.
